# A head-to-head comparison of multiple amyloid PET radiotracers for Down syndrome clinical trials

**DOI:** 10.1101/2025.03.18.25324200

**Authors:** Matthew D. Zammit, Julie C. Price, Bradley T. Christian, Michael S. Rafii, Alzheimer’s Biomarker Consortium – Down Syndrome

## Abstract

Adults with Down syndrome carry high risk of developing Alzheimer’s disease and efforts to include this population in clinical trials remain limited. A barrier to recruitment for anti-amyloid trials includes the availability of the same amyloid PET radiotracer to multiple treatment centers. 237 adults with Down syndrome from the Trial-Ready Cohort – Down syndrome and Alzheimer Biomarker Consortium – Down syndrome studies were imaged using T1-w MRI and using PET images of PiB, Florbetapir (FBP), NAV4694 (NAV) or Flutemetamol (FMM) to screen for amyloid plaque (Aβ) burden. PiB displayed the largest effect size to measure amyloid change while FBP had a small effect size. NAV and PiB, which are structurally similar compounds, displayed similar sensitivity to measure longitudinal amyloid increase. The estimated age at amyloid onset showed no significant difference between PiB, FBP, NAV or FMM. The findings suggest that different amyloid PET radiotracers provide consistent estimates of amyloid onset age for adults with Down syndrome. Multicenter studies of Alzheimer’s disease therapeutics can utilize multiple amyloid PET radiotracers to facilitate recruitment, however, PiB and NAV displayed the greatest sensitivity to detect longitudinal change.

## Introduction

Individuals with Down syndrome (DS) represent the largest population of genetically determined Alzheimer’s disease (AD)^1^. This genetic form of AD is driven by triplication of chromosome 21, which encodes the gene responsible for amyloid precursor protein production, leading to beta-amyloid plaque (Aβ) deposition early in life. AD is accountable for 70% of deaths in this demographic over the age of 35^1–3^. The prevalence of AD dementia sharply increases past 50 years of age and the average age of dementia onset is 55 years^4^. As the lifetime risk of AD in DS exceeds 90%^5,6^, DS may serve as a model population for understanding both the progression of the disease and the long-term benefits of early therapeutic interventions. Clinical trials in neurotypical populations during early-stage dementia have shown the success of anti-amyloid therapies at removing Aβ and slowing the progression of clinical symptoms^7^. AD biomarker studies suggest these drugs may be more effective for secondary prevention by administering during the “preclinical” timeframe before clinical symptoms emerge. Despite the severity of AD risk in DS, this population has not been included in recent anti-amyloid clinical trials; resulting from factors such as limited access to expert clinical evaluation for dementia, challenges in assessing cognitive change in the setting of intellectual disability, concerns regarding cerebral amyloid angiopathy, and the presumed increased risk of adverse events related to amyloid-related imaging abnormalities (ARIA) common with these therapies^1,8^.

Amyloid plaque pathology can be detected with PET imaging and amyloid burden increases with age^9^. Anti-amyloid therapeutic trials utilize PET outcome measures to confirm target engagement and rely on radiotracers labeled with fluorine-18 that allow for regional distribution to treatment centers. Many large DS imaging studies have historically used carbon-11 labeled Pittsburgh Compound B (PiB), which cannot be distributed in a multicenter trial due to its short half-life, and a knowledge gap exists on how fluorine-18 radiotracers evaluate longitudinal Aβ change with respect to AD staging in this population. To address these limitations and to improve the feasibility of DS recruitment for clinical trials, we present the first multi-radiotracer comparison for longitudinal Aβ change from the Trial Ready Cohort – Down syndrome (TRC-DS) and the Alzheimer Biomarker Consortium – Down syndrome (ABC-DS).

## Materials and methods

A total of 237 adults with DS were recruited from the TRC-DS and ABC-DS studies. Participants from each cohort were included in the study if they had available and quality checked MRI and amyloid PET. Institutional Review Board approval and informed consent were obtained during enrollment into the study by the participant or legally designated caregiver according to the Declaration of Helsinki. Participant demographics and imaging site information are provided in Table 1. All participants underwent T1-w MRI imaging, as well as PET imaging using either PiB, Florbetapir (FBP), NAV4694 (NAV) or Flutemetamol (FMM) across multiple study visits (Table 1). Global Aβ burden was determined using the Centiloid (CL) method^10^ using the whole cerebellum as the reference tissue for PiB, NAV and FMM, while an eroded white matter reference was used for FBP. Briefly, PET images were spatially normalized to MNI152 template space, standardized uptake value ratio (SUVR) images were then generated by voxel normalization to the mean activity in the reference tissue, and the mean cortical SUVR was extracted from the Centiloid cortex ROI. Cortical SUVR was then converted to units of CL using the appropriate conversion equations for PiB^10^, FBP^11^, NAV^12^ and FMM^13^. Participants were classified as amyloid-positive (A+) using an a priori threshold of 18 CL^14^. Using a sampled iterative local approximation (SILA) algorithm^15^, a longitudinal CL trajectory model was created for the DS population as described previously from the ABC-DS^14,16^. Briefly, the participant’s PET CL outcomes were fit to the model to determine their ‘Amyloid Age,’ or the estimated number of years since becoming PET A+. Amyloid Age was then subtracted from the participants Chronological Age at their most recent scan to determine their estimated age at amyloid onset.

**Table 1.**
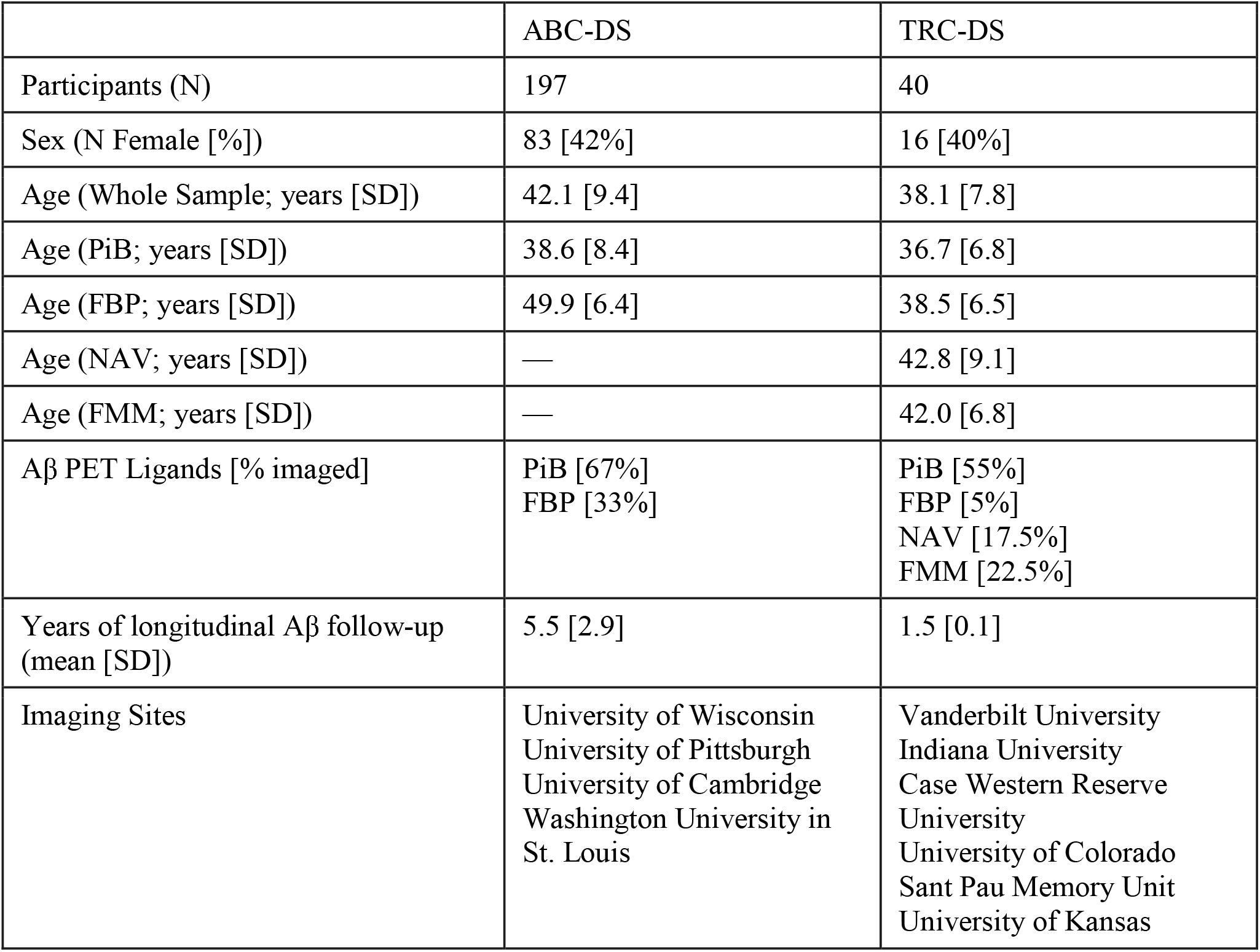
Participant demographics from ABC-DS and TRC-DS.

|  | ABC-DS | TRC-DS |
| --- | --- | --- |
| Participants (N) | 197 | 40 |
| Sex (N Female [%]) | 83 [42%] | 16 [40%] |
| Age (Whole Sample; years [SD]) | 42.1 [9.4] | 38.1 [7.8] |
| Age (PiB; years [SD]) | 38.6 [8.4] | 36.7 [6.8] |
| Age (FBP; years [SD]) | 49.9 [6.4] | 38.5 [6.5] |
| Age (NAV; years [SD]) | — | 42.8 [9.1] |
| Age (FMM; years [SD]) | — | 42.0 [6.8] |
| Aβ PET Ligands [% imaged] | PiB [67%]<br>FBP [33%] | PiB [55%]<br>FBP [5%]<br>NAV [17.5%]<br>FMM [22.5%] |
| Years of longitudinal Aβ follow-up (mean [SD]) | 5.5 [2.9] | 1.5 [0.1] |
| Imaging Sites | University of Wisconsin<br>University of Pittsburgh<br>University of Cambridge<br>Washington University in St. Louis | Vanderbilt University<br>Indiana University<br>Case Western Reserve University<br>University of Colorado<br>Sant Pau Memory Unit<br>University of Kansas |

## Statistical analyses

Rates of longitudinal Aβ change were measured in CL/year for each radiotracer, and Cohen’s d effect sizes^17^ were calculated to evaluate the differences in radiotracer binding between the baseline and most recent follow-up measure. Estimated amyloid onset ages were then compared between the radiotracers using one-way ANOVA with Tukey’s HSD for multiple comparisons. An alpha of *P* < .05 was used to determine statistical significance.

## Results

Amyloid PET outcomes using PiB, FBP and NAV displayed longitudinal increases with respect to both Chronological Age and Amyloid Age in the TRC-DS and ABC-DS cohorts (Figure 1). Longitudinal FMM data was not obtained during this study. Table 2 provides longitudinal outcomes for each radiotracer as well as the estimated amyloid onset ages determined for these cohorts. For participants imaged with PiB, the mean baseline amyloid burden was 40 [18] CL while the follow-up amyloid burden was 58 [22] CL, corresponding to an effect size (Cohen’s d) of 0.9. Participants scanned with FBP displayed mean baseline amyloid burden of 70 [33] CL, and follow-up burden of 80 [37] CL, corresponding to an effect size of 0.3. Only one participant received longitudinal NAV imaging, and this individual had a baseline and follow-up amyloid burden of 44 and 51 CL, respectively. The mean baseline amyloid burden for all participants scanned with NAV was 41.5 [2.2] CL, while the mean baseline burden for FMM was 65.5 [5.2] CL. Longitudinal rates of amyloid change were calculated as 7.3 [4.1], 3.3 [3.5] and 4.9 [0.0] CL/year for PiB, FBP and NAV, respectively. From ANOVA, participants scanned with PiB displayed significantly higher rates of amyloid increase compared to FBP [F(df) = 8.2(1); *P* = .007]. Following temporal modeling to align the participants’ trajectories to the population-averaged Amyloid Age curve, the estimated age at A+ onset was determined as 41.9 [6.4] years for PiB, 42.9 [6.0] years for FBP, 43.2 [4.7] years for NAV and 47.0 [9.1] years for FMM. From ANOVA, no significant differences in estimated age at A+ onset were observed between radiotracers (F(df) = 1.4(1); *P* = .23). Multiple comparison analysis with Tukey’s HSD revealed no significant differences in estimated age at A+ between each radiotracer.

**Table 2.** Amyloid onset age, amyloid PET burden, the rate of longitudinal amyloid increase, and Cohen’s d effect sizes for participants with Down syndrome scanned with PiB, FBP, NAV and FMM. Amyloid burden is presented in units of Centiloids (CL).

| PET Radiotracer | Estimated Amyloid Onset Age (years [SD]) | A+ Baseline Amyloid Burden (CL [SD]) | A+ Follow-up Amyloid Burden (CL [SD]) | A+ Longitudinal rate of change (CL/year [SD]) | Cohen's d Effect Size |
| --- | --- | --- | --- | --- | --- |
| PiB | 41.9 [6.4] | 40 [18] | 58 [22] | 7.3 [4.1] | 0.9 |
| FBP | 42.9 [6.0] | 70 [33] | 80 [37] | 3.3 [3.5] | 0.3 |
| NAV | 43.2 [4.7] | 41.5 [2.2] | 51 [0.0]* | 4.9 [0.0]* | N/A* |
| FMM | 47.0 [9.1] | 65.5 [5.2] | N/A** | N/A** | N/A** |

\*Only one participant had longitudinal NAV scans
\*\*No longitudinal FMM scans were performed during this study

**Figure 1.**
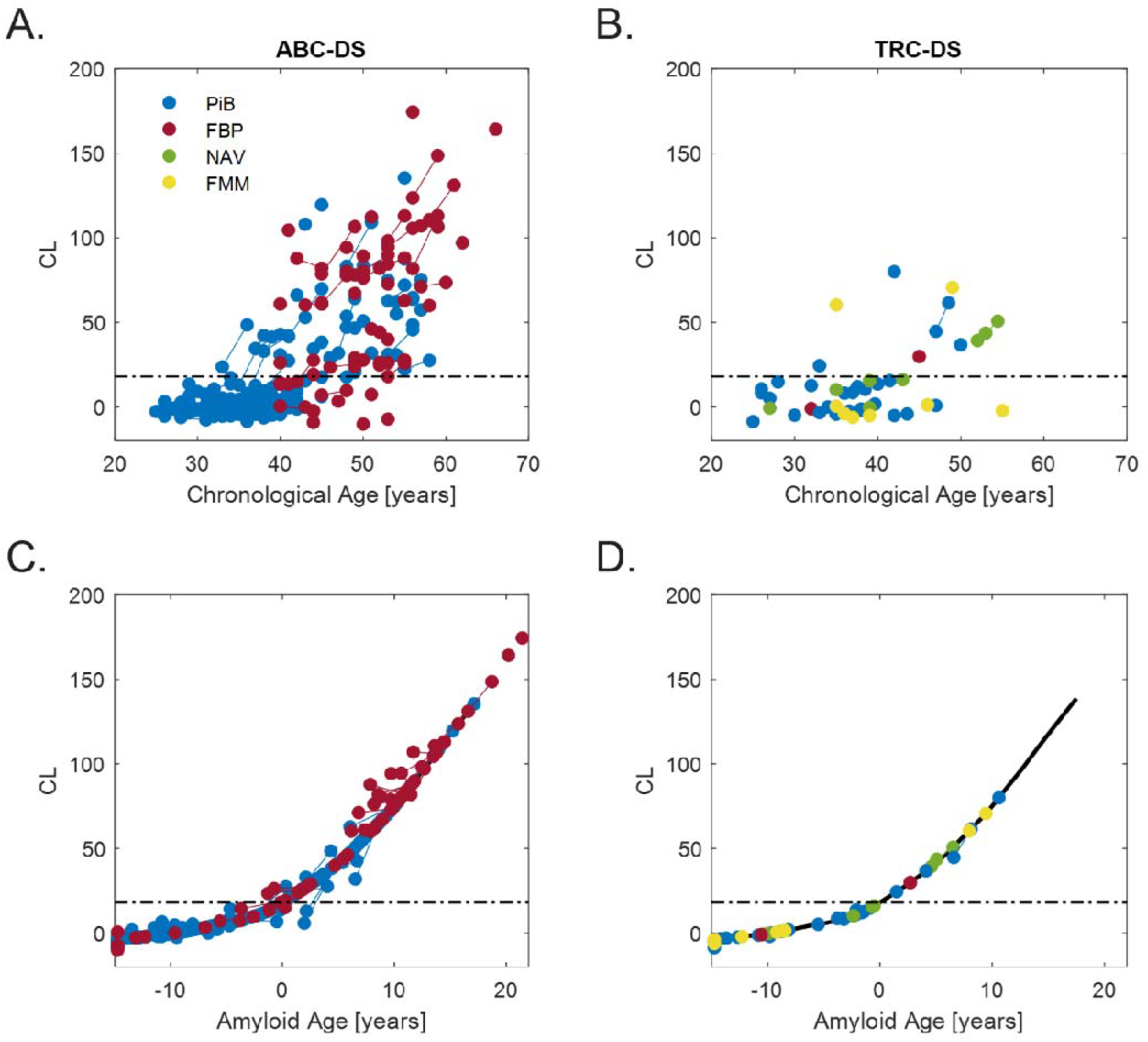
Cross-sectional (single point) and longitudinal (points connected by lines) amyloid trajectories for ABC-DS and TRC-DS. A. Amyloid PET Centiloids (CL) with respect to Chronological Age for ABC-DS participants evaluated with PIB and FBP. **B**. Amyloid PET CL with respect to Chronological Age for TRC-DS participants evaluated with PIB, FBP, NAV and FMM. **C**. Modeled longitudinal trajectories displaying CL with respect to Amyloid Age in ABC-DS. **D**. Modeled longitudinal trajectories displaying CL with respect to Amyloid Age in TRC-DS. Dashed lines represent the a priori threshold for A+ of 18 CL. The solid black curve in C&D represents the previously established CL vs. Amyloid Age trajectory for the Down syndrome population.

## Discussion

The low prevalence of DS (∼12.6 in 10000 births) in the United States^18^ poses a challenge for clinical trial recruitment, especially within the age range where AD-related change begins. This study represents the first multi-radiotracer comparison of amyloid change for the DS population to facilitate DS trial recruitment. This study also represents a large, multicenter project to collect AD biomarker data and stage the DS population for disease-modifying clinical trials. Historically, ABC-DS published amyloid imaging findings using PiB PET, and this new data provides comparisons of PiB PET to FBP, NAV and FMM with the addition of the TRC-DS dataset. When evaluating the amyloid PET findings at the cross-sectional level, all four radiotracers were able to provide similar estimates in the age at amyloid onset. This suggests that use of any amyloid PET radiotracer should be suitable for the purposes of staging an individual within the AD continuum and for the purposes of clinical trial screening. For longitudinal analysis, PiB, FBP and NAV were able to detect longitudinal amyloid increase between baseline and follow-up measures, with TRC-DS having the shortest follow-up of ∼1.5 years, which aligns with the timeline of anti-amyloid trial endpoints. Longitudinal imaging data for FMM as part of TRC-DS was not available for the current study. Longitudinal FMM data will be collected as the TRC-DS study progresses. PiB displayed the greatest effect size to detect longitudinal increase, while FBP displayed the smallest effect size, likely a result of its lower signal dynamic range^11^. In the current study, we chose to use an eroded white matter reference region for FBP as studies suggest improved quantitative accuracy over the cerebellum^19^. However, the sample of adults scanned with FBP were in an age range where cognitive impairment is more prevalent, yet the sensitivity of FBP to detect Aβ change was much lower than the sensitivity of PiB measured in a younger cohort, suggesting poor longitudinal performance of FBP. While limited in sample size, NAV displayed greater longitudinal increase compared to FBP. NAV results could be anticipated to be closely aligned with PiB, as binding and in vivo kinetics have been shown to be highly correlated with PiB while displaying lower off-target white matter signal^20^ compared to FBP. NAV and PiB are structurally similar^12^, resulting in similar in vivo performance.

Previous AD-related studies in the DS population highlight the heterogeneity in the age at amyloid onset^21,22^. Thus, clinical trial recruitment based upon age alone would result in significantly high screening fail rates to achieve a suitable sample size. A recent study observing the prevalence of Aβ positivity identified that only 5% of individuals between the ages of 35-39 were A+^9^. This prevalence increased to ∼57% between the ages of 40-44. In DS it has been observed that both neurofibrillary tau deposition^14^ and cognitive decline^23,24^ occur rapidly following the onset of amyloid, with some estimates showing change as early as 5 years after PET A+ is reached^16^. The rapid onset of tau and cognitive decline suggests that a narrow window of opportunity exists to treat AD in DS, highlighting that age-based clinical trial recruitment alone is not sufficient. Use of biomarker data as part of the screening process can help facilitate recruitment by minimizing screening fail rates. In this study, we incorporated use of a sampled iterative local approximation (SILA) algorithm^15^ to align the PET outcomes of our participants to a DSAD specific timeline of amyloid accumulation. This methodology has been performed previously in DS^16^, allowing for a robust characterization of the progression of AD following Aβ onset. This amyloid clock^25^ framework has the potential to aid clinical trial design for DS as the AD therapeutic field moves towards implementing combination therapies targeting multiple AD pathologies. In the current study, we highlight that individuals with DS can be evaluated within the amyloid clock using multiple amyloid PET radiotracers which display similar estimates in the age at amyloid onset. Taken together, use of multiple amyloid PET radiotracers can help facilitate clinical trial recruitment in this at-risk population.

The findings suggest that the amyloid PET radiotracers PiB, Florbetapir, NAV4694 and Flutemetamol provide consistent estimates of amyloid onset age for adults with Down syndrome. Multicenter studies of Alzheimer’s disease therapeutics can utilize multiple amyloid PET radiotracers to facilitate recruitment, however, PiB and NAV displayed the greatest sensitivity to detect longitudinal Aβ change.

## Abbreviations

(Aβ): beta-amyloid
(PET): positron emission tomography
(PiB): Pittsburgh compound B
(FBP): Florbetapir
(NAV): NAV4694
(FMM): Flutemetamol
(AD): Alzheimer’s disease
(DS): Down syndrome
(CL): Centiloid
(SUVR): Standardized uptake value ratio

## Data Availability

The imaging sites have entered web-based data through the Alzheimer’s Therapeutic Research Institute (ATRI) as part of the ABC-DS study. The data that support the findings of this study are openly available in the ABC-DS database (https://ida.loni.usc.edu/login.jsp?project=ABCDS).

## Acknowledgements

Alzheimer’s Biomarker Consortium–Down Syndrome (ABC-DS) Investigators:

Beau M. Ances, MD PhD; Howard F. Andrews, PhD; Karen Bell, MD; Rasmus M. Birn, PhD; Adam M. Brickman, PhD; Peter Bulova, MD; Jeff Burns, MD; Amrita Cheema, PhD; Kewei Chen, PhD; Bradley T. Christian, PhD; Isabel Clare, PhD; Ann D. Cohen, PhD; Eric W. Doran, MS; Tatiana M. Foroud, PhD; Benjamin L. Handen, PhD; Jordan Harp, PhD; Sigan L. Hartley, PhD; Elizabeth Head, PhD; Denise Head, PhD; Christy Hom, PhD; Lawrence Honig, MD; Milos D. Ikonomovic, MD; Sterling C Johnson, PhD; M. Ilyas Kamboh, PhD; David Keator, PhD; Julia K. Kofler, MD; William Charles Kreisl, MD; Sharon J. Krinsky-McHale, PhD; Florence Lai, MD; Patrick Lao, PhD; Charles Laymon, PhD; Joseph Hyungwoo Lee, PhD; Ira T. Lott, MD; Victoria Lupson, PhD; Mark Mapstone, PhD; Davneet Singh Minhas, PhD; Neelesh Nadkarni, MD; Sid O’Bryant, PhD; Deborah Pang, MPH; Melissa Petersen, PhD; Julie C. Price, PhD; Lauren Ptomey, PhD; Margaret Pulsifer, PhD; Michael S. Rafii, MD PhD; Herminia Diana Rosas, MD; Frederick Schmitt, PhD; Nicole Schupf, PhD; Wayne P. Silverman, PhD; Dana L. Tudorascu, PhD; Rameshwari Tumuluru, MD; Badri Varadarajan, PhD; Michael A. Yassa, PhD; Shahid Zaman, MD PhD; Fan Zhang, PhD

## Author contributions

MDZ conceptualized the project, performed data processing and analysis and drafted the manuscript. JCP performed data processing and analysis and edited the manuscript. BTC edited the manuscript. MSR conceptualized the project and edited the manuscript.

